# Impact of anti-androgen therapies on COVID-19 susceptibility: a case-control study in male population from two COVID-19 regional centers of Lombardy (Italy)

**DOI:** 10.1101/2020.04.20.20068056

**Authors:** Massimo Lazzeri, Stefano Duga, Elena Azzolini, Vittorio Fasulo, Nicolò Buffi, Alberto Saita, The Humanitas COVID-19 Task Force, The Humanitas Gavazzeni COVID-19 Task Force, Rodolfo Hurle, Alessandro Nobili, Maurizio Cecconi, Paolo Casale, Rosanna Asselta

**Author notes:** Equally contributed (Massimo Lazzeri, Stefano Duga). **Corresponding Authors:** Massimo Lazzeri, MD, PhD, Department of Urology, Istituto Clinico Humanitas IRCCS, Via Manzoni 56 (20089) Rozzano – Milan (Italy), Stefano Duga, PhD, Department of Biological Sciences, Humanitas University, Via Rita Levi Montalcini 4 (20090) Pieve Emanuele - Milan (Italy).

## Abstract

**Importance:** There are gender differences in vulnerability to the Coronavirus disease 2019 (COVID-19), with men experiencing more severe disease at any age. The use of anti-androgen drugs, like 5-alpha reductase inhibitors (5ARIs), could protect from severe pulmonary disease.

**Objective:** To determine whether men who received 5ARIs for benign prostatic hyperplasia (BPH) have a lower risk of hospitalization for COVID-19.

**Design:** This is a case-control study on patients hospitalized for COVID-19 (cases), matched to beneficiaries of the Lombardy Regional Health Service (controls).

**Setting:** Data were collected by two high-volume COVID-19 regional centres of Lombardy (Italy) from 1^st^ March to 24^th^ April 2020.

**Participants:** Consecutive patients positive for SARS-CoV-2 virus according to the WHO guidance, who required hospitalization.

**Exposure:** BPH treatment with 5ARIs (finasteride/dutasteride) in the last six months.

**Main Outcome(s) and Measure(s):** The primary outcome was to compare the prevalence of male patients chronically exposed to 5ARIs, who required hospitalization for COVID-19, with the one of age-matched males in Lombardy.

**Results:** Overall, 1,432 COVID-19 patients were included. Among the 943 males, 45 (4.77%) patients were exposed to chronic 5ARI therapy. COVID-19 patients aged >55 years under 5ARI treatment were significantly less than expected on the basis of the prevalence of 5ARI treatment among age-matched controls (5.57 vs. 8.14%; p=0.0083, 95%CI=0.75-3.97%). This disproportion was even higher for men aged >65 (7.14 vs. 12.31%; p=0.0001, 95%CI=2.83-6.97%).

Eighteen 5ARIs-patients died; the mean age of men who died was higher than those who did not: 75.98±9.29 vs. 64.78±13.57 (p<0.001). Cox regression and multivariable models did not show any correlation between the exposure to 5ARIs and protection against ICU admission or death: HR=0.79 (95%CI=0.54-1.15; p=0.22) and OR=1.23 (95%CI=0.81-1.87; p=0.33), respectively.

**Conclusions and Relevance:** In this case-control study the use of 5ARIs was less frequent among patients hospitalized for COVID-19 than among controls, suggesting that men exposed to 5ARIs might be less vulnerable to severe COVID-19. This observation further supports the idea to test in randomized clinical trials whether anti-androgen therapies can prevent the transition from paucisymptomatic SARS-CoV-2 infection to overt pulmonary disease.

**KEY POINTS:** *Question:* May treatment of benign prostatic hyperplasia with 5-alpha reductase inhibitors (5ARIs) impact on COVID-19-related hospitalization risk?

*Findings:* In this case-control study, which compared 943 adult males hospitalized for COVID-19 with age-matched men from Lombardy (all beneficiaries of the Regional Health Service), the proportion of patients aged >55 years, who were exposed to 5ARIs (dutasteride, finasteride), was significantly lower (5.57%) than that of the general male population (8.14%) (p=0.0083, 95%CI=0.75-3.97%).

*Meaning:* The use of 5ARIs was associated with a potential reduced risk of hospitalization for COVID-19 in men older than >55 years. This suggests the opportunity to test whether anti-androgen therapies can prevent the transition from paucisymptomatic SARS-CoV-2 infection to overt pulmonary disease.

## MAIN TEXT

Over the past months, the Coronavirus disease 2019 (COVID-19) pandemic has marched relentlessly westward. Presently, Italy has one of the highest rates of SARS-CoV-2 infection (363.100,000 people), and one of the highest mortality rates, 13.98% vs. an average value of 6.75% (as of May 11th, 2020 [1]). Although sex-disaggregated data for COVID-19 in Italy show equal numbers of cases between men and women [https://www.epicentro.iss.it/coronavirus/], there seem to be gender differences in vulnerability to the disease, with men more prone to have higher severity and lethality, independently of age (male.female lethality ratio=1.78). Similar data were found in almost all populations, with male.female lethality ratios up to 3.2 (mean 1.62±0.45) [https://globalhealth5050.org/covid19/] [2]. Experience from past outbreaks shows the importance of incorporating a gender analysis into preparedness and response efforts to improve the effectiveness of health interventions and treatments [3]. The spike (S) protein of coronaviruses mediates viral entry into target cells [4]. Entry depends on binding of the surface unit, S1, of the S protein to angiotensin converting enzyme 2 (ACE2). Moreover, entry requires S-protein priming by cellular proteases, which allows fusion of viral and cellular membranes, and is mediated mainly by the cellular serine protease TMPRSS2 [5]. TMPRSS2 gene expression is responsive to androgen stimulation [6] and it was hypothesized that differences in TMPRSS2 expression levels by androgen route might be implicated in the observed sex differences [7]. Moreover, the hormonal differences between males and females may impact on a number of genes and pathways, and can vary according to age and reproductive status [8]. Therefore, a treatment reducing androgen stimulation might eventually impact on disease severity by different mechanisms.5-alpha reductase inhibitors (5ARIs), used for the treatment of benign prostatic hyperplasia (BPH) and alopecia, inhibit the 5-alpha reductase enzyme, responsible for the conversion of testosterone into the more active dihydrotestosterone [9]. Importantly, all three isoforms of the 5-alpha reductase (SRD5A1-3) enzyme are expressed in the lung [https://gtexportal.org/home].

We hence tested the hypothesis that men who are chronically exposed to 5ARIs for BPH treatment could have a lower risk of hospitalization for COVID-19 and different outcomes, compared to men who don’t. The primary outcome was to compare the frequency of 5ARIs treatment among male patients hospitalized for COVID-19 with the drug-prescription rate among sex-matched controls (as inferred from drug-prescription data of 2018 of the Lombardy Regional Health Service, LRHS). The secondary outcome was to determine disease severity according to hospitalization days, access to intensive care unit (ICU), and outcome at discharge (alive vs. death) in 5ARI-treated COVID-19 patients vs. non-treated ones.

This is an observational case-control study on consecutive patients admitted to two COVID-19 centers of Lombardy (Italy): Istituto Clinico Humanitas IRCCS, Milan and Humanitas Gavazzeni, Bergamo, from March 1^st^ to April 24^th^, 2020, who were positive for SARS-CoV-2 virus according to WHO guidances. The study was approved by the Ethics Committee of the Istituto Clinico Humanitas on March 16^th^, 2020 (protocol number 233/20). SARS-CoV-2 laboratory confirmation was defined as a positive result of real-time RT-PCR assay on rhino-pharyngeal swabs or bronchoalveolar lavage. A complete medical/pharmacological history was obtained for each patient (by himself, relatives, his general practitioner). We focused on men who have been taking 5ARIs (finasteride, dutasteride) in the last six months (chronic exposure) for BPH treatment. Descriptive statistical analysis was performed. Hazard ratios (HR), odds ratios (OR), and 95% confidence intervals (CI) for associations between 5ARI treatment and outcomes were estimated by regression models.

To test the hypothesis that exposure to 5ARIs may affect the risk to be hospitalized for COVID-19, patients were stratified according to the following age classes: 45-54, 55-64, 65-74, 75-84 and ≥85 years. For each class, the prevalence of 5ARI treatment for BPH (single or in combination with alpha blockers, AB) was compared to the one of the general male Lombardy population, as obtained from LRHS (whose organization is described in [10]).

Data from 1,432 COVID-19 patients were analyzed (489 females, 943 males); their demographic/clinical characteristics are summarized in Supplementary Table 1, whereas those specifically referring to males are summarized in Tables 1-2. Overall, 45 (4.77%) among male patients were exposed to a chronic 5ARI therapy; 16 of them received a therapy with an AB too. Eight patients (0.85%) suffered from PCa and were treated with androgen deprivation therapy (ADT).

**Table 1:** Summary of COVID-19 male patients (demographics, comorbidities, and BPH/PCa therapies).

| *Characteristic* | *Level* | *Alive* | *Dead* | *P value* |
| --- | --- | --- | --- | --- |
| **N** |  | 662 | 281 |  |
| **Age, mean (±SD)** |  | 64.78 (13.57) | 75.98 (9.29) | **<0.001** * |
| **Hospitalizations days, mean (±SD)** |  | 14.83 (9.96) | 8.60 (6.89) | **<0.001** * |
| **ICU** | No | 545 (82.33%) | 233 (82.92%) | 0.74 § |
|  | Yes | 117 (17.67%) | 47 (16.73%) |  |
| **Day in ICU, mean (±SD)** |  | 11 (9.89) | 8.38 (6.72) | **<0.001** * |
| **Comorbidity** | No | 285 (43.05%) | 84 (29.89%) | **<0.001** § |
|  | Yes | 377 (56.95%) | 197 (70.11%) |  |
| **Age categories** | 55-64 | 166 (25.08%) | 22 (7.83%) | **<0.001** § |
|  | 65-74 | 181 (27.34%) | 74 (26.33%) |  |
|  | 75-84 | 131 (19.79%) | 132 (46.98%) |  |
|  | Others § | 195 (29.46%) | 56 (19.93%) |  |
| **Therapy for BPH or PCa** | No therapy \$\$ | 587 (88.67%) | 231 (82.21%) | |
|  | 5ARIs | 16 (2.42%) | 13 (4.63%) | 0.053 § |
|  | 5ARIs + Alpha-Blocker | 11 (1.66%) | 5 (1.78%) | 0.79 § |
|  | ADT | 5 (0.75%) | 2 (0.71%) | 0.98 § |
|  | Alpha-Blocker | 43 (6.49%) | 29 (10.32%) | **0.031** § |
|  | ADT + Alpha-Blocker | 0 (0%) | 1 (0.36%) | 0.11 § |

\* = Student's t test.
§ = Pearson's chi-square test.
\$ = The category "others" includes all subjects aged <55 years and ≥ 85 years
\$\$ = used as reference category in the Pearson's chi-square test.
ADT = Androgen deprivation therapy; BPH = Benign prostatic hyperplasia; ICU = Intensive care unit; PCa = Prostate cancer; SD = Standard deviation.
Significant P value are in bold.

**Table 2:**
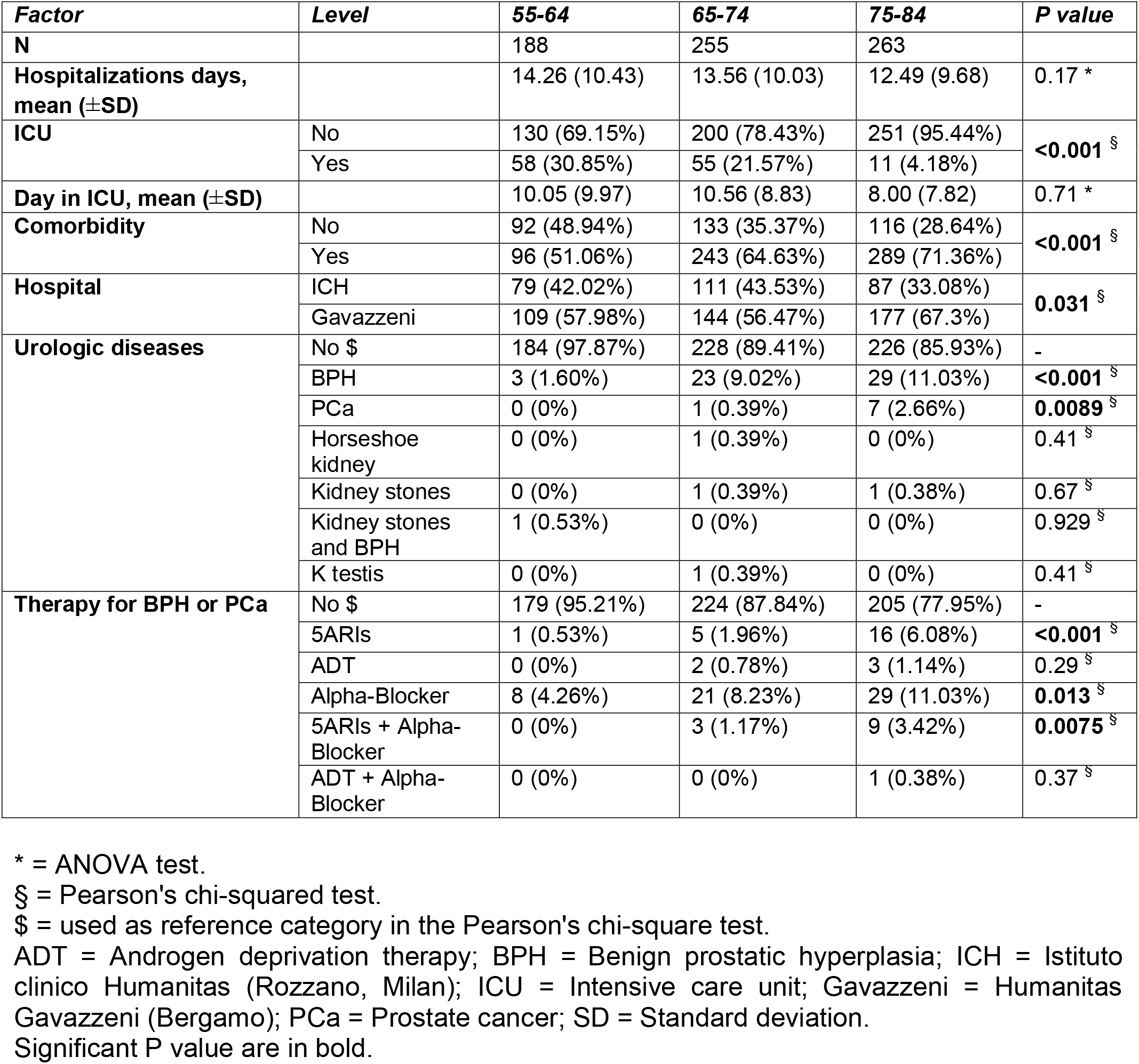
COVID-19 male patient characteristics according to age.

Among the 45 patients under 5ARIs, 18 died; the mean age of men who died was not significantly different for the one of survivors: 78.39±6.82 vs. 79.15±10.46 (p=0.78). The overall lethality rate for males taking 5ARIs was 40%, higher than the one calculated for all male patients. This can be explained by the higher age of males taking 5ARIs (Tables 1, 2): 78.84±9.10 vs. 68.12±13.45 (p<0.001). Finally, Cox regression model and multivariable analyses did not show any correlation between the exposure to 5ARIs and protection against ICU admission or death: HR=0.79 (95%CI=0.54-1.15; p=0.22) and OR=1.23 (95%CI=0.81-1.87; p=0.33), respectively.

Table 3 shows that 5ARIs were less frequently prescribed in COVID-19 patients than in the general population. The percentage of patients >55 years who received 5ARIs was 5.57% vs. 8.14% (p=0.0083, 95%CI=0.75-3.97%). In particular, age classes 65-74 and 75-84 showed a percentage of 5ARI prescriptions of 3.14% and 9.51%, among hospitalized patients vs. 7.90% and 16.06% among controls (relative differences, 4.76% and 6.55%; p=0.0048 and 0.0038, respectively).

**Table 3:** Impact of anti-androgen therapies on COVID-19.

|  | **LOMBARDY MALE POPULATION** |  |  | **THIS STUDY – COVID-19 PATIENTS** |  |  |  |  |
| --- | --- | --- | --- | --- | --- | --- | --- | --- |
| **Age range** | **Males in Lombardy \*** | **Patients treated with 5ARIs** | **Patients treated with 5ARIs or 5ARIs (%)** | **Males in Hospitals** | **Patients treated with 5ARIs or 5ARIs+AB (%)** | **P value (95%CI)** | **P value \*\* (95%CI)** | **P value \*\*\* (95%CI)** |
| 45-54 **** | 847062 | 1827 | 0.22 | 101 | 0 (0) | 0.64<br>(-3.44-0.23) |  |  |
| 55-64 | 657301 | 11930 | 1.81 | 188 | 1 (0.53) | 0.19<br>(-1.14-1.72%) | **0.0083**<br>**(0.75-3.97%)** |  |
| 65-74 | 520850 | 41152 | 7.90 | 255 | 8 (3.14) | **0.0048**<br>**(1.83-6.30%)** |  | **0.0001**<br>**(2.83-6.97%)** |
| 75-84 | 361967 | 58136 | 16.06 | 263 | 25 (9.51) | **0.0038**<br>**(2.40-9.54%)** |  |  |
| ≥85 | 115170 | 23555 | 20.45 | 84 | 10 (11.90) | 0.052<br>(-0.092-13.86%) |  |  |

\* The here-reported male population of Lombardy does not include individuals living in nursing houses, being their pharmacological prescriptions not included in the Lombardy Regional Health Service database.
\*\* Considering merged age categories 55-64, 65-74, 75-84, and ≥85.
\*\*\* Considering merged age categories 65-74, 75-84, and ≥85.
\*\*\*\* For completeness also
Chi-square analysis was used for comparisons of proportions.
CI = Confidence interval.

We showed that the proportion of COVID-19 patients aged >55 years under 5ARI treatment was significantly lower than expected on the basis of the prevalence of 5ARI treatment among controls of the same age range. Concerning 5ARI prescription, estimates are to be considered "conservative”, because LRHS reports do not include boxes bought in pharmacies without asking for reimbursement from the National Health Service, a tendency more likely among younger patients, also considering the affordable costs of generic drugs.

Our study has some limitations: i) it is a descriptive observational study in a highly selected population within a short-time window; ii) though we adopted the most recent release, the dataset from the Lombardy region used for comparisons date back to 2018; iii) the sample size is relatively small for an epidemiological survey targeting a large population; iv) the sample almost exclusively consists of Caucasians, so the study cannot be generalized to other ethnic groups. In this regard, it is interesting to note that African Americans, who are more susceptible to COVID-19 (infection and death rates for predominantly black counties in USA is >3-fold and 6-fold higher than in predominantly white counties) [11], also have higher PCa rates and poorer prognosis [12].

The present study provides preliminary evidence that the use of 5ARIs might represent a protective factor against hospitalization for COVID-19 among men aged >55 years. This effect might be even stronger in men under ADT, as androgen levels would be zero. Our cohort is not enough powered to evaluate the effect of ADT, which would be interesting to assess on larger datasets. In this respect, promising data were recently released by Montopoli and colleagues [13], who evidenced that cancer patients have an increased risk of SARS-CoV-2 infections compared to non-cancer patients, with PCa patients receiving ADT being instead partially protected from infections.

Among possible spin offs of our results is the possibility to test anti-androgen drugs in COVID-19 patients to prevent severe lung disease. Should this be possible with 5ARIs instead of ADT, we could avoid the adverse events associated with total castration (libido loss, cardiovascular diseases, osteoporosis, sarcopenia).

## Data Availability

not applicable

## ACKNOWLEDGMENTS

**Members of the Humanitas COVID-19 Task Force:** Aghemo Alessio, Angelini Claudio, Badalamenti Salvatore, Balzarini Luca, Bocciolone Monica, Cecconi Maurizio, Ciccarelli Michele, Kurihara Hayato, Lagioia Michele, Omodei Paolo, Preatoni Paoletta, Voza Antonio, Accornero Stefano, Alfarone Ludovico, Ali Hussam, Arcari Ivan, Arosio Paola, Azzolini Elena, Baccarin Alessandra, Baggio Sara, Barbagallo Michela, Barberi Caterina, Barbic Franca, Barbieri Viviana, Barbone Alessandro, Basciu Alessio, Benvenuti Chiara, Bianchi Ilaria, Borea Federica, Borroni Mario, Bresciani Gianluigi, Brunetta Enrico, Bulletti Cinzia, Cadonati Cristina, Calabro’ Loenzo, Calatroni Marta, Calvetta Albania Antonietta, Cannata Francesco, Canziani Lorenzo, Capogreco Antonio, Capretti Giovanni Luigi, Carlani Elisa, Carrone Flaminia, Casana Maddalena, Ceribelli Angela, Ceriotti Carlo, Cimino Matteo, Ciuffini Leonardo, Colaizzi Chiara, Colapietro Francesca, Costa Guido, Cozzi Ottavia, Craviotto Vincenzo, Crespi Chiara, Crippa Massimo, Da Rio Leonardo, Dal Farra Sara, D’antonio Federica, De Ambroggi Guido, De Donato Massimo, De Lucia Francesca, De Nittis Pasquale, De Santis Maria, Delle Rose Giacomo, Di Pilla Marina, Dipaola Franca, Dipasquale Andrea, Dipasquale Angelo, Droandi Ginevra, Fazio Roberta, Ferrante Giuseppe, Ferrara Elisa Chiara, Ferrari Matteo Carlo, Ferri Sebastian, Folci Marco, Foresti Sara, Franchi Eloisa, Fraolini Elia, Furfaro Federica, Galimberti Paola, Galtieri Alessia, Gavazzi Francesca, Generali Elena, Goletti Benedetta, Guidelli Giacomo, Jacobs Flavia, Lania Andrea Gherardo, Libre’ Luca, Lleo Ana, Loiacono Ferdinando, Lughezzani Giovanni, Maccallini Marta, Magnoni Paola, Maiorino Alfonso Francesco, Malesci Alberto, Mantovani Riccardo, Marchettini Davide, Marinello Arianna, Markopoulos Nikolaos, Masetti Chiara, Mazziotti Gherardo, Milani Angelo, Mirani Marco, Morelli Paola, Motta Francesca, Mundula Valeria, Nigro Mattia, Ormas Monica, Pagliaro Arianna, Paliotti Roberta, Pavesi Alessia, Pedale Rosa, Pegoraro Francesco, Pellegatta Gaia, Pellegrino Marta, Petriello Gennaro, Piccini Sara, Pocaterra Daria, Poliani Laura, Procopio Fabio, Puggioni Francesca, Pugliese Luca, Pugliese Nicola, Racca Francesca, Randazzo Michele, Regazzoli Lancini Damiano, Reggiani Francesco, Rodolfi Stefano, Ruongo Lidia, Sacco Clara, Sagasta Michele, Sandri Maria Teresa, Savi Marzia, Scarfo’ Iside, Shiffer Dana, Sicoli Federico, Solano Simone, Solitano Virginia, Stainer Anna, Stella Matteo Carlo, Strangio Giuseppe, Taormina Antonio, Testoni Lucia, Tordato Federica, Trabucco Angela, Ulian Luisa, Valentino Rossella, Valeriano Chiara, Vena Walter, Verlingieri Simona, Vespa Edoardo, Zanuso Valentina, Zilli Alessandra, Lutman Fabio, Lanza Ezio, Profili Manuel, Giannitto Caterina, Mrakic Sposta Federica, Torrisi Chiara, Poretti Dario, Pedicini Vittorio, Bonifacio Cristiana, D’orazio Federico, D’antuono Felice, Castelli Alice, Pestalozza Alessandra, Paiardi Silvia, Teofilo Francesca Ilaria, Citterio Gianluigi, Aloise Monia, Ripoll Pons Marta, Lavezzi Elisabetta, Fedeli Carlo, Desai Antonio, Caltagirone Giuseppe, Voza Giuseppe, Giorgino Massimo Giovanni.

**Members of the Humanitas Gavazzeni COVID-19 Task Force**. Abati Elena Maria; Abualkheir Mohammed; Agradi Sergio; Albano Giovanni; Alioto Giuseppina; Allegrini Davide; Altieri Vincenzo Maria; Angeli Enzo; Arduini Mario; Assisi Alberto; Baldi Marzia; Barbagallo Lucia; Barzaghi Maria Elena; Basili Luca Manfredi; Beretta Alessandra; Beretta Natascia; Beretta Giordano; Beretta Simone; Bertella Erika; Bettoni Elisabetta; Biazzo Alessio; Bombardieri Emilio; Bonacina Manuela; Bonfanti Riccardo; Bordoni Mariagrazia; Bordoni Luca; Borghesi Maria Lucia; Bortolotti Luigi; Bravi Marco; Brena Federica; Bruno Simona; Camozzini Valentina; Canziani Lorenzo Maria; Caporarello Salvatore; Cappelleri Gianluca; Caputo Rosilde; Carrara Alfonso; Carriero Federica; Castoldi Massimo; Castriota Fausto; Catellani Francesco; Catenacci Alberto; Celotti Simona; Cereda Marco Angelo; Ceresoli Giovanni Luca; Cesari Eugenio; Chiesa Giuseppe; Colli Alessandro; Corapi Antonio; Cordua Nadia; Cortina Gabriele; Coscione Andrea Vittorio; Costa Maria Concetta; Cremonesi Alberto; Dalto Serena; D’Aquino Nicola; D’Aveni Alessandro; De Amicis Francesco; De Filippis Costantino Nicandro; Delalio Elena; Dell’Era Valentina; Di Cintio Davide; Di Noia Vincenzo Pio; Esposito Giovanni; Fedele Isabella; Ferrari Elisa; Filippi Claudia; Finamora Ilaria; Fiorentino Gennaro; Fortino Olga; Franceschini Grisolia Enrico; Frongillo Elisabettamaria; Fumagalli Miriam; Gabuda Marian Svyatoslavovich; Gaffuri Nicola; Galbussera Maurizio; Galdamez Carcamo Shintia Coralia; Galli Sara; Gentinetta Franco; Gerometta Piersilvio; Ghilardi Carlo Guardo; Ghirardelli Paolo; Ghirardi Guido Luciano; Ghisi Patrizia Chiara; Giambartino Sebastiano; Giofre’ Fabrizio; Giroletti Laura; Goletti Orlando; Graniero Ascanio; Grassi Massimo Maria; Grazioli Valentina; Guanella Giovanni Battista; Guidotti Eugenio; Intelisano Antonio; Lanzone Alberto Maria; Ledda Giovanna Franca; Licini Gloria; Liguori Alessia; Lisanti Rocca Carmela; Lleshaj Eliona; Loffreno Antonella; Lopresti Ennio; Lucca Elena; Macca Claudio; Macchini Daniele; Maggi Lamberto; Magni Luca; Mancin Annalisa; Manfredini Fabio; Marangoni Silvia; Marchese Stefano; Marzano Bernardo; Mascioli Giosue’; Masia Antonio Francesco; Mazzocchi Claudia; Mazzoni Maurizio Giovanni; Meco Massimo; Mela Federico; Meroni Roberta; Micciche’ Eligio; Mingone Daniela; Molteni Mattia; Monti Cinzia; Moretti Antonio; Nerla Roberto; Nicoletti Alessandro; Nicoli Flavia; Orlandi Roberto; Paleologo Claudia; Passaretti Bruno Maria; Pedrigi Maria Cristina; Pesenti Nicola; Polidoro Roberto; Poloni Camillo Luca; Previtali Ilaria; Quartierini Giorgio; Rabbolini Giovanni; Re Chiara; Rea Bruna; Ricciardelli Gabriella; Rizzardi Giovanna; Ronzoni Annalisa; Roscitano Claudio; Rossi Daniela; Rota Stefano; Ruggiero Perrino Vincenzo; Salvini Piermario; Santoro Franco; Sauta Maria Grazia; Schifilliti Daniela; Scrofani Amalia; Selmi Carlo; Setti Lucia; Smarrelli Davide; Solinas Costantino; Spreafico Andrea Giovanni; Squadroni Michela; Stagno Maria Francesca; Stelian Edmond Jean; Stratta Gregorio; Tarantino Luca; Tessa Lorenzo; Testa Amidio; Testa Rosa Miranda; Traini Mariaemilia; Trovati Serena; Uccelli Fara Margerita Letizia; Usai Luca; Valenti Francesco; Vandenbulcke Filippo; Vavassori Vittorio Luigi; Vernile Laura; Viganò Luca; Villa Clara; Villa Giambattista; Villari Nicola; Vismara Alberto Carlo; Zambarbieri Giulia; Zanello Alessandro; Zerbini Graziano; Zotti Mariacamilla; Zumbo Aurora.

We also thank Dr. Mauro Tettamanti for helping us in retrieving data from the Lombardy Regional Health Service.

This work was supported by Ricerca Corrente (Italian Ministry of Health), intramural funding (Fondazione Humanitas per la Ricerca). Generous contributions of the Banca Intesa San Paolo and of Dolce&Gabbana Fashion Firm are gratefully acknowledged.

